# Simulating the impact of vaccination rates on the initial stages of a COVID-19 outbreak in New Zealand (Aotearoa) with a stochastic model

**DOI:** 10.1101/2021.11.22.21266721

**Authors:** Leighton M. Watson

## Abstract

**Aim:** The August 2021 COVID-19 outbreak in Auckland has caused the New Zealand government to transition from an elimination strategy to suppression, which relies heavily on high vaccination rates in the population. As restrictions are eased and as COVID-19 leaks through the Auckland boundary, there is a need to understand how different levels of vaccination will impact the initial stages of COVID-19 outbreaks that are seeded around the country.

**Method:** A stochastic branching process model is used to simulate the initial spread of a COVID-19 outbreak for different vaccination rates.

**Results:** High vaccination rates are effective at minimizing the number of infections and hospitalizations. Increasing vaccination rates from 20% (approximate value at the start of the August 2021 outbreak) to 80% (approximate proposed target) of the total population can reduce the median number of infections that occur within the first four weeks of an outbreak from 1011 to 14 (25^th^ and 75^th^ quantiles of 545-1602 and 2-32 for V=20% and V=80%, respectively). As the vaccination rate increases, the number of breakthrough infections (infections in fully vaccinated individuals) and hospitalizations of vaccinated individuals increases. Unvaccinated individuals, however, are 3.3x more likely to be infected with COVID-19 and 25x more likely to be hospitalized.

**Conclusion:** This work demonstrates the importance of vaccination in protecting individuals from COVID-19, preventing high caseloads, and minimizing the number of hospitalizations and hence limiting the pressure on the healthcare system.

## Introduction

In response to the COVID-19 global pandemic, New Zealand initially followed an elimination strategy that coupled tough lockdowns with strict border controls. The first COVID-19 case was reported on 28 February 2020 and on 19 March 2020 international borders were closed to all but New Zealand citizens and permanent residents.^1^ On 25 March 2020, New Zealand moved to Alert Level 4 (level 4 is the toughest restrictions in the four-level alert system) with strict lockdown measures including the closure of educational and public facilities along with all non-essential businesses, stay-at-home orders, limits on travel, and no gatherings allowed. From 10 April 2020, anyone entering the country had to undergo two weeks of managed isolation and quarantine (MIQ). These measures kept case numbers low with a total of 1504 cases before COVID-19 was declared eliminated in New Zealand on 8 June 2020.^1^

Mandatory quarantine of overseas arrivals has been broadly effective at keeping COVID-19 out of New Zealand. In the period up to 15 June 2021 there have been 10 border-related re-incursions,^2^ although these outbreaks were able to be quickly detected and successfully eliminated. On 17 August 2021, a COVID-19 case of the highly transmissible delta variant with no clear link to the border was detected in Auckland. As a result, the entire country moved from Alert Level 1 (minimal restrictions) to Alert Level 4 (toughest restrictions). The lockdown measures and the implementation of an internal boundary around Auckland have been largely successful in containing the outbreak to Auckland. However, at the time of writing cases have been detected in Northland and Waikato with a limited number of further cases detected around the country, including in the South Island. After 35 days at Alert Level 4 in Auckland, the government began to ease restrictions and transition from an elimination to suppression strategy.

On 22 October 2021, the government outlined the COVID-19 Protection Framework, which uses vaccination certificates along with public health measures to manage COVID-19 in the community. This system will replace the Alert Level framework when District Health Boards reach a vaccination target of 90% of the eligible population. As vaccination rates increase and restrictions are eased, it is expected that COVID-19 will spread out of Auckland to other parts of the country (as has already been observed with outbreaks in Waikato and Northland). It is important to understand the impact of different vaccination rates on the growth of future outbreaks outside of Auckland along with the number of hospitalizations and hence stress on the healthcare system.

In this work a stochastic branching process model^3-5^ is used to simulate the initial stages of a COVID-19 outbreak within a community. Stochastic models are useful because they incorporate the randomness associated with the initial stages of an outbreak. For some simulations, COVID-19 will spread widely and form an extensive outbreak (e.g., Auckland in August 2021 where a returnee from Australia sparked an outbreak with >6000 cases at the time of writing) whereas for other simulations, despite the virus having a reproduction number larger than one, random chance will mean that COVID-19 does not spread far beyond the initial seed infection (e.g., Wellington in June 2021 when a COVID-19 infected traveller visited from Australia but did not infect anyone else). The stochastic model tracks each individual case and becomes computationally expensive for large case numbers. Therefore, while stochastic models are useful for simulating the initial stages of an outbreak, deterministic SEIR (Susceptible, Exposed, Infected, Recovered) models are frequently used for larger and longer-term population level studies of epidemics.^5,6^

Here, I use a stochastic model to study the how the number of infections and hospitalizations depend on the vaccination rate and population level controls. I calculate the likelihood that a new infected case or hospitalized individual is vaccinated and determine the relative risk of getting infected or hospitalized with COVID-19 between vaccinated and unvaccinated individuals.

## Methods

The stochastic model presented here tracks the number of infections in the community and categorizes individuals as symptomatic (clinical infections) or asymptomatic (subclinical infections). Each infected individual infects a random number of other individuals, *N*, drawn from a Poisson distribution (Figure 1)^3^. For a symptomatic individual, the Poisson distribution is defined by *λ = RC* where *R* is the reproduction number and *C* is the effectiveness of population level controls (e.g., Level 1, 2, 3, or 4 in the Alert Level Framework or Green, Orange, or Red in the COVID-19 Protection Framework). For an asymptomatic individual, the Poisson distribution is defined by *λ = RC/2*, which assumes that asymptomatic individuals infect, on average, half as many people as symptomatic individuals.^7^

**Figure 1:**
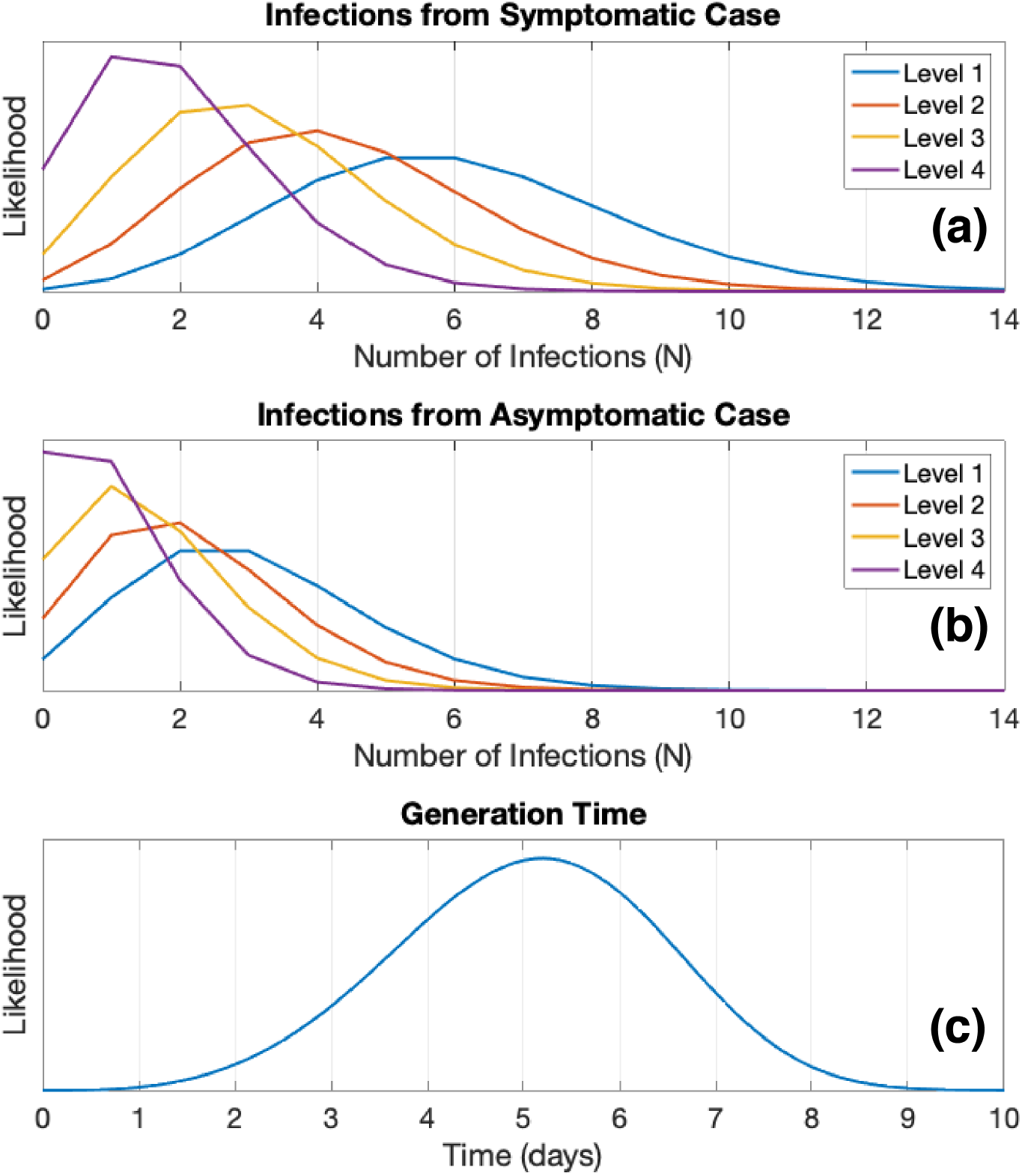
Probability distributions used in the stochastic model. (a) Number of infections caused by a symptomatic case and (b) by an asymptomatic case. The number of infections caused by a symptomatic or asymptomatic case is governed by a Poisson distribution and can only take integer values. (c) The generation time, which is the time between an individual getting infected and infecting others, is governed by a Weibull distribution.

Population level controls include public health measures such as physical distancing, wearing of masks, closure of schools and non-essential businesses, and restrictions on gatherings and social activities. The effectiveness of population level controls are taken from Plank et al.^3^ as *C*=1 for Alert Level 1, *C*=0.72 for Alert Level 2, *C*=0.52 for Alert Level 3, and *C*=0.32 for Alert Level 4. These values were estimated for the initial variant of COVID-19 and population level controls may be less effective against the more transmissible delta variant.^8^ In addition, the model does not account for illegal gatherings or other non-compliance with restrictions.

The generation times between an individual becoming infected and infecting *N* other individuals are independently sampled from a Weibull distribution with *a*=5.57 and *b*=4.08 where *a* is the scale parameter and *b* is the shape parameter (mean=5.05 days and variance=1.94 days)^9^ (Figure 1). The model assumes that 33% of new infections are asymptomatic (subclinical) with the remainder symptomatic (clinical).^10-12^

I consider a range of vaccination rates, *V*, from 0% to 90% of the total population (rather than the *eligible population*, which at the time of writing is the over 12 years old population). Unlike previous work by Steyn et al.^5,13^ age is not accounted for in the model, either in the vaccination rollout where older individuals are more likely to be vaccinated, or in the susceptibility where older individuals are more likely to experience severe disease or death. The vaccination rate *V* is assumed to be constant throughout the simulated outbreak. Following Steyn et al.^5,13^ the Pfizer-BioNTech vaccine, which is the only COVID-19 vaccine currently being widely administered in New Zealand, is assumed to be 70% effective against infection and 50% effective against transmission for breakthrough infections.^14^ Throughout the simulations, the model tracks the total number of vaccinated and unvaccinated infections along with the number of symptomatic and asymptomatic cases. Infected individuals are assumed to be equally likely to interact with vaccinated and unvaccinated individuals, with probabilities based solely on the vaccination rate. This may lead to an underestimation of the spread of COVID-19 in unvaccinated communities as unvaccinated individuals are more likely to have unvaccinated contracts.

The likelihood of hospitalizations is also modeled. Clinical infections have a 7.8% probability of being hospitalized.^15^ This agrees with the total number of cases hospitalized during the August 2021 Auckland outbreak (7.6% hospitalization rate for all cases as of 30 October 2021).^16,17^ Based on Dagan et al.^18^ who examined the effectiveness of the Pfizer-BioNTech vaccine against hospitalization in Israel, it is assumed that the vaccine is 87% effective at preventing hospitalizations after two doses. The model only allows for cases to be fully vaccinated (defined as more than two weeks after the second dose of the two-dose Pfizer-BioNTech vaccine) or unvaccinated. The model does not include the additional complexity of individuals who are partially vaccinated, either by only receiving one dose of the vaccine or by being within two weeks after receiving the second dose. In addition, there is no lag time between becoming infection and becoming hospitalized. The model is only run for a short duration (28 days) and hence I do not simulate the likelihood of hospitalized individuals dying.

Note that the model presented here does not include any testing, contact tracing, or isolation of cases. Instead, I focus on the impact of vaccination rates, particularly on the early stages of an outbreak when cases may be circulating undetected. The reader is referred to Steyn et al.^5^ for a model that includes testing and estimates the number of infections at the time of detection of the outbreak for various vaccination rates and testing scenarios.

## Results

The simulations are seeded with one unvaccinated symptomatic individual at *t*=0 where *t* is the time in days. Simulations are run for 28 days with time steps of 1 day. The model tracks each infected individual, distinguishes between symptomatic cases (clinical infections) and asymptomatic cases (subclinical infections), tracks hospitalization rates, and distinguishes between vaccinated and unvaccinated individuals. I consider vaccination rates between 0% and 90% of the total population in 5% increments (the vaccination rate referred to here is the total population rather than the eligible population, which is over 12 years old at the time of writing). The government’s vaccination target of 90% of the eligible population corresponds to 78.7% of the total population.^16,17^ For each vaccination rate, we consider four different population level controls based on New Zealand’s Alert Level system using the effectiveness values from Plank et al.^3^ To get a representative sample of the possible outcomes for each scenario, we run the model 100,000 times for each combination of vaccination rate and population level controls.

### Vaccination Rate and Population Level Controls

I consider the impact of the vaccination rate and population level controls on the total number of infections and hospitalizations. Figure 2 shows likelihood of the number of infections 28 days into an outbreak for V=0%, 30%, 60%, and 90% and no population level controls (*C=1*). The histograms indicate the likelihood of each number of infections and illustrate the randomness associated with the initial stages of an outbreak. For some simulations, random chance causes the outbreak to infect a small number of people whereas for other simulations the outbreak can rapidly grow due to super-spreader events. For V=30%, there is a 10% chance that an outbreak will cause less than 98 infections after 28 days. However, there is also a 10% chance that an outbreak will cause more than 1340 infections. The potential spread of an outbreak is strongly dependent on the vaccination rate. For V=0%, there is a 50% chance than an outbreak will cause less than 2691 infections after 28 days compared to less than 75 infections for V=60%.

**Figure 2:**
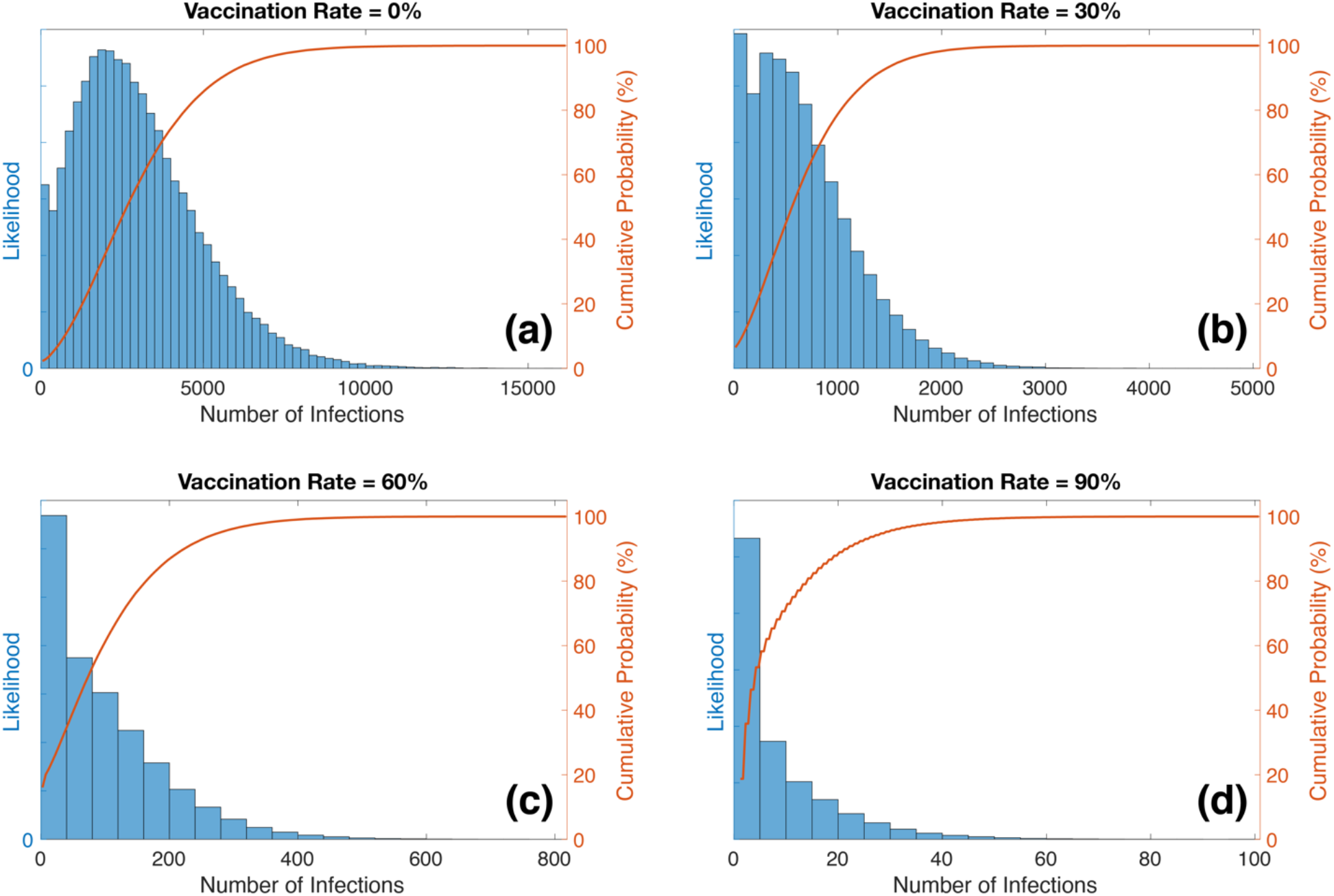
Histograms showing the likelihood for a given number of infections 28 days into an outbreak for vaccination rates of (a) 0%, (b) 30%, (c) 60%, and (d) 90%. Red lines indicate the cumulative probability.

The number of infections is strongly dependent on the vaccination rate with higher vaccination rates decreasing the number of infections by several orders of magnitude. For V=0%, the maximum number of infections after 28 days is 16,198 compared to 819 for V=60% and 101 for V=90%. The strong dependence of the number of infections on the vaccination rate is further visualized in Figure 3, which shows the median number of infections along with the 25% and 75% quartiles as a function of vaccination rate and population level controls. Increasing the vaccination rate can drastically decrease the number of infections and hence minimize the size of an outbreak.

**Figure 3:**
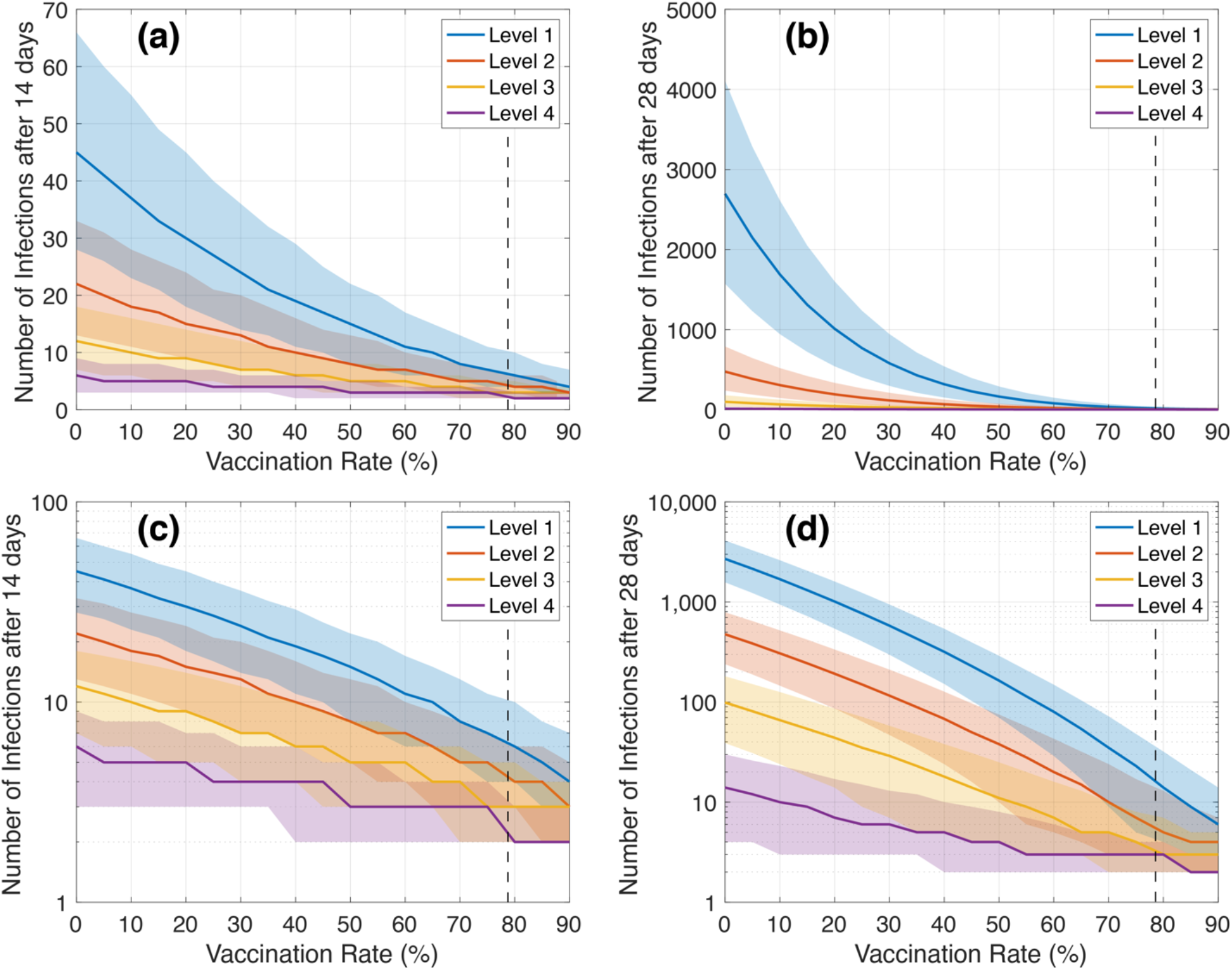
Number of infections as a function of vaccination rate and population level controls for (a,c) 14 days and (b,d) 28 days after an unvaccinated symptomatic individual is seeded into the community. Solid lines show the median of the 100,000 realizations while the shaded area shows the range between the 25% and 75% quantiles. Top plots (a,b) show the results on a linear scale while the bottom plots (c,d) show the results on a logarithmic scale. The vertical black dashed lines indicate vaccination rate for the total population that corresponds to the 90% target of the over 12 years old population (78.7%).

Figure 3 also shows the importance of population level controls, especially at low levels of vaccination. For V=20% (the approximate vaccination rate in New Zealand on 17 August 2021 when the delta outbreak was first detected), the median number of infections after 28 days is 1,011 (542 and 1602 for 25% quantile and 75% quantile, respectively) at Level 1 but only 7 (3 and 17 and for 25% quantile and 75% quantile, respectively) at Level 4. This illustrates how, in the absence of high vaccination rates, population level controls are extremely important in limiting the growth of an outbreak. It is noted that New Zealand only shifted from Level 1 to Level 4 after a case was detected. At this point, the virus had been circulating undetected for approximately a week prior to this and had already seeded 800 to 1000 cases in the community.^19^ Furthermore, the effectiveness values used here for the population level controls were estimated for the original strain of Covid-19.^3^ It is unclear if these values are appropriate for the more transmissible delta variant^8,20,21^ or if population level controls are less effective against delta.

The results shown in Figures 2 and 3 illustrate how a vaccine can be extremely effective in preventing or limiting an outbreak even if the vaccine does not provide individuals with 100% protection from infection. Here, it is assumed that the Pfizer-BioNTech vaccine is 70% effective in preventing infection and, for breakthrough infections, 50% effective at preventing onward transmission.^14^ Despite this imperfect protection, at a population level the vaccine drastically reduces the spread of the virus. For V=60%, the mean number of infections 28 days into the outbreak is only 3% of the mean number of infections for V=0%. For V=90%, the mean number of infections further decreases to only 0.3% compared to the unvaccinated scenario.

The number of hospitalizations is also strongly dependent on the vaccination rate (Figure 4). For V=0%, there is a 50% chance of over 282 hospitalizations 28 days into the outbreak compared to 50% chance of less than 6 hospitalizations for V=60% and 1 hospitalization for V=90%. The hospitalization results presented in Figure 4 are calculated for C=1, which gives a worst-case scenario. Implementation of population level controls (i.e., Alert Level restrictions or vaccination certificates) will reduce the total number of hospitalizations at all vaccination rates but the general trend is unchanged; high vaccination rates drastically reduce the number of hospitalizations.

**Figure 4:**
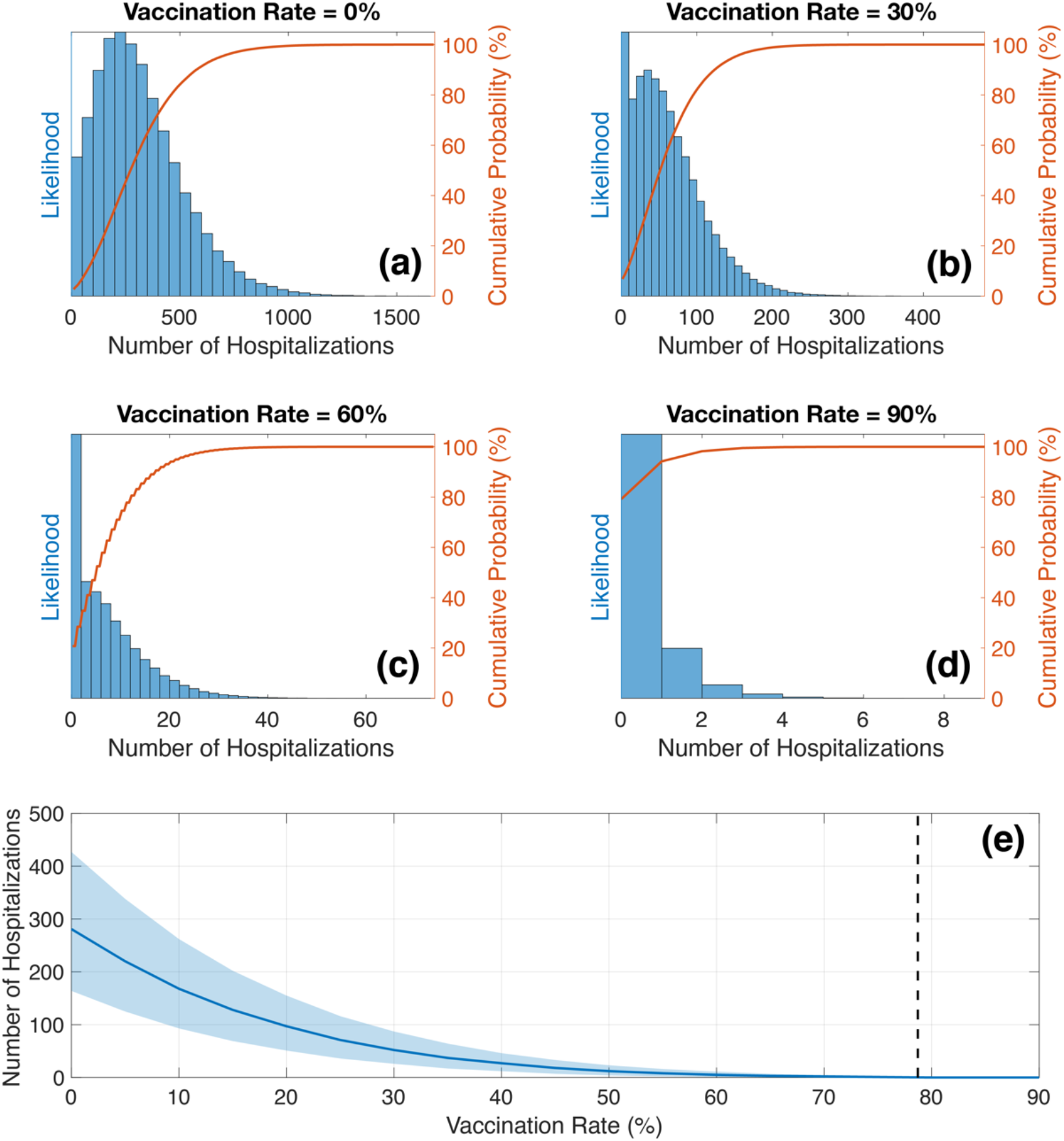
Histograms showing the likelihood for a given number of hospitalizations 28 days into an outbreak for C=1 and vaccination rates of (a) 0%, (b) 30%, (c) 60%, and (d) 90%. Red lines indicate the cumulative probability. (e) Number of hospitalizations 28 days into an outbreak as a function of vaccination rate showing the (solid line) median simulation result and (shaded area) 25^th^ and 75^th^ quantiles. Vertical black line indicates the vaccination rate of the total population that corresponds to 90% of the eligible (over 12 years old) population (78.7%).

### Risk factor for unvaccinated individuals

As the vaccination rate increases, there are more vaccinated individuals in the population, fewer unvaccinated individuals, and hence more breakthrough infections will occur (infections in fully vaccinated individuals). This is expected; the model assumes a 30% chance of breakthrough infections in vaccinated individuals.^14^ As the number of vaccinated individuals increases, the number of cases in vaccinated individuals will increase, as shown in Figure 5a. For V=40%, there is an 83% chance that a new case will be unvaccinated and a 17% chance that they will be vaccinated. For V=80%, this switches to 46% unvaccinated and 54% vaccinated. It is important to note that, even though the number of infections in vaccinated individuals increases, it stays below the population proportion, which is shown by the dashed lines in Figure 5a. This indicates that, per population, infections are still more likely to occur in unvaccinated individuals. It should also be noted that infections in unvaccinated individuals are more likely to lead to onward transmission and hospitalization than breakthrough infections.^14,18^

**Figure 5:**
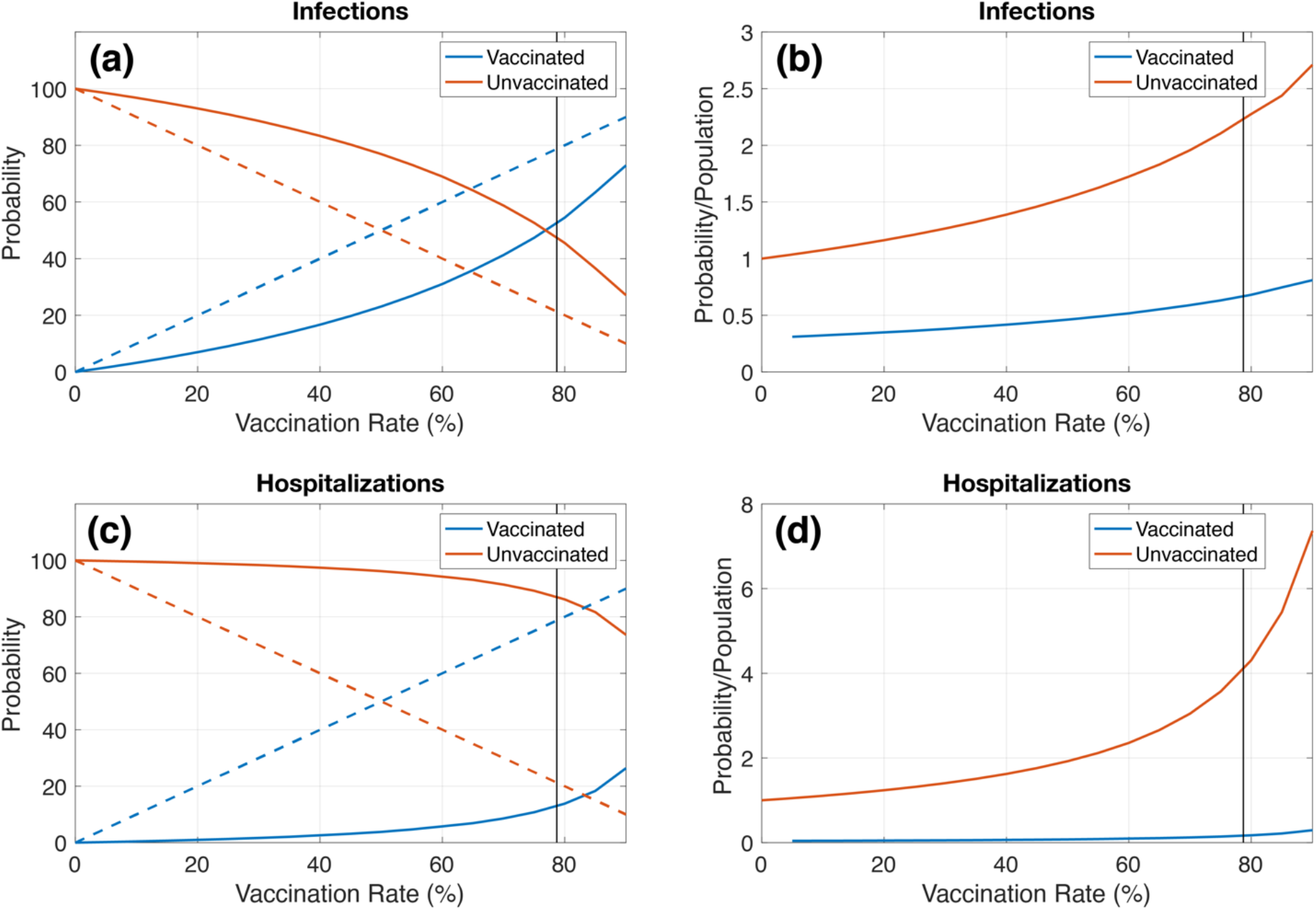
Probability that an infected or hospitalized individual is (blue) vaccinated or (red unvaccinated as a function of vaccination rate, as calculated from mean simulation result. (a) and (c) Probability that an (a) infected or (c) hospitalized individual is vaccinated or unvaccinated. Dashed lines indicate proportion of population that are vaccinated or unvaccinated as a function of vaccination rate. (b) and (d) Probability normalized by population that an (b) infected or (d) hospitalized individual is vaccinated or unvaccinated.

Figure 5b shows the probability that an infection occurs in a vaccinated or unvaccinated individual normalized by population. At all vaccination rates, an unvaccinated individual is more likely to be infected than vaccinated individual. Dividing the unvaccinated probability normalized by population by the vaccinated probability normalized by population gives the risk factor for unvaccinated individuals, which quantifies the likelihood of an unvaccinated individual getting infected compared to a vaccinated individual. This shows that, even though the number of breakthrough infections increases with increasing vaccination rate, an unvaccinated individual is 3.3x more likely to be infected than a vaccinated individual for all vaccination rates.

I then perform the same analysis for hospitalizations. Figure 5c shows that the probability that a hospitalized case is vaccinated increases slower than the probability that an infected case is vaccinated. This shows that while the vaccine provides substantial protection against infection, it provides even greater protection against hospitalization. At all vaccination rates, hospitalizations are significantly more likely to be unvaccinated than vaccinated (at V=80%, there is an 86% chance that a hospitalized case will be unvaccinated compared to 14% vaccinated). Figure 5d shows the probability of hospitalization normalized by population. Unvaccinated individuals are 25x more at risk of hospitalization than vaccinated individuals. This reinforces the need to vaccinate a large percent of the population to minimize hospitalizations and prevent strain on the healthcare system.

The results shown here are in broad agreement with a CDC study of 43,127 COVID-19 cases in Los Angeles County that showed that infection and hospitalization rates among unvaccinated individuals were 4.9x and 29.2x, respectively, higher than for fully vaccinated individuals.^22^ The risk factors calculated here may be lower due to underestimating the protection that the vaccine provides against infection or because I focus on the delta variant, which transitioned to becoming the dominant variant during the study period of Griffin et al.^22^

## Conclusions

The current COVID-19 outbreak in New Zealand, which began in August 2021, has resulted in the highest case counts experienced in New Zealand during the pandemic. Cases are centered in Auckland but have spread to Northland and Waikato, with isolated cases spread around the rest of the country, including the South Island. The government’s switch from an elimination to suppression strategy has emphasized the importance of vaccination in preventing COVID-19 from overwhelming the healthcare system. As restrictions ease, there is a need to understand how different vaccination rates will impact the initial stages of COVID-19 outbreak as cases become seeded in communities around New Zealand.

Here, I use a stochastic branching process model to examine the impact of vaccination rates on the initial spread of an outbreak. I show that increasing vaccination rates greatly decrease the number of infections (1.4% median number of infections 28 days into the outbreak for V=80% compared to V=20%), even if the Pfizer-BioNTech vaccine only provides individuals with imperfect protection (assumed to be 70% effective against breakthrough infection). This illustrates the effectiveness of the vaccine on a population level.

As the vaccination rate increases, the number of breakthrough infections and hospitalizations among vaccinated individuals will increase. This is expected and reflects the increased proportion of vaccinated individuals in the population. Unvaccinated individuals are 3.3x more likely to be infected and 25x more likely to be hospitalized than vaccinated individuals. The model results presented here agree with real-world data^22^ and highlight how the Pfizer-BioNTech vaccine provides good protection against infection and extremely good protection against hospitalization. This work illustrates the need for high vaccination rates to reduce infections and prevent the healthcare system from being overrun with COVID-19 patients.

## Data Availability

The code used in the present study are available upon reasonable request to the author

